# Predicting Olanzapine Induced BMI increase using Machine Learning on population-based Electronic Health Records

**DOI:** 10.1101/2025.08.26.25334441

**Authors:** Foteini D. Aktypi, Michael E. Benros, Simon Rasmussen

## Abstract

**Background:** Weight gain is a common side effect in patients treated with olanzapine (N05AH03), contributing to increased risks of metabolic complications such as diabetes, cardiovascular disease, and reduced treatment adherence. However, personalised prevention strategies are currently lacking in day-to-day clinical practice. Identifying factors that can predict which patients are most likely to gain a significant amount of weight is therefore critical. Such insights could enable early risk prediction and guide clinicians in implementing personalized interventions to minimize side effects and improve long-term treatment outcomes.

**Methods:** In this study, we developed explainable machine learning models using population-based Electronic Health Record data and compared the performance between logistic regression, decision trees and ensemble tree classifiers like XGboost to predict significant BMI increase of >5% in the next contact to the clinic while on olanzapine.

**Results:** XGBoost model achieved the highest performance for predicting a >5% BMI increase on olanzapine with an AUROC of 0.72, surpassing logistic regression (AUROC = 0.69) and other evaluated classifiers. Model performance was consistent across sexes, but varied across age groups, with the highest performance for individuals 30-69 years of age (AUROC = 0.73) and lowest in individuals over 70 years of age (AUROC = 0.67). The SHAP analysis highlighted several key predictive features including prolonged intervals between follow-up visits, higher baseline BMI, younger age, shorter time since olanzapine initiation, cumulative hospitalization days, increasing olanzapine dosage, polypharmacy and especially concurrent use of sedatives as well as multiple prescriptions with anxiolytics.

**Conclusion:** Our models validate prior known risk factors for olanzapine induced weight gain and further uncover previously unknown factors that influence >5% BMI increase on olanzapine. These findings underscore the need for continued research in this domain to establish effective preventive measures for individuals undergoing antipsychotic treatments.

## Introduction

Antipsychotic medications are primarily used for treatment of psychotic disorders such as schizophrenia, but some antipsychotics can also be used as mood stabilizers for bipolar disorder, and as adjunctive therapies for major depressive disorder. Among the antipsychotics the second-generation antipsychotics (SGAs), also known as atypical antipsychotics, have been increasingly used due to their lower risk of neurological side effects compared to first-generation antipsychotics (Leucht et al., 2013). Olanzapine, an SGA introduced in the late 1990s, has become one of the most prescribed antipsychotics due to its efficacy in managing psychotic symptoms and its mood-stabilizing properties (Beasley et al., 1997). Pharmacologically, olanzapine acts as an antagonist at dopamine D2 and serotonin 5-HT2A receptors while also modulating other neurotransmitter pathways (Ceskova, 2020).

Despite its therapeutic benefits, olanzapine is frequently implicated in significant metabolic side effects, through weight gain (Tschoner et al., 2007). The consequences of weight gain include increased risk of metabolic syndrome, characterized by central obesity, dyslipidemia, hypertension, and insulin resistance, which substantially elevates the risk of cardiovascular disease and type 2 diabetes (DE Hert et al., 2009; Newcomer, 2007). Medication-induced weight gain also affects adherence to treatment, due to patient concerns about their appearance and overall health, which increases the likelihood of hospitalizations, posing a significant economic burden (Gianfrancesco et al., 2006; Riordan et al., 2011). It is widely reported that a significant proportion of individuals experience weight gain exceeding 5% of baseline body weight, with olanzapine consistently ranking among the top contributors to these increases with an average weight increase of 3-8 kg in a period of 6-12 months (Bazo-Alvarez et al., 2020; Huang et al., 2020; Huhn et al., 2019). Meta-analyses also show that weight gain could exceed 7 kg within a year of treatment, particularly in younger patients (Citrome et al., 2011; Nasrallah, 2008).

Modeling the Body Mass Index (BMI), a measure calculated as weight in kilograms divided by the square of height in meters used to estimate body fat and assess health risks, presents unique challenges due to the irregular and sparse nature of BMI measurements in routine clinical practice of Electronic Health Record (EHR) data. Patients usually have their weight recorded only sporadically during appointments or hospitalizations, making it difficult to capture dynamic BMI trends in a systematic manner. Furthermore, patient heterogeneity, arising from differences in genetics, baseline metabolic health, and lifestyle factors, introduces unknown confounding variables that complicate predictive modeling since, this is not routinely collected information (Miotto et al., 2018).

Past studies on olanzapine and BMI have explored predictive models to understand and manage weight gain. Previous models have shown promise in identifying individuals at greater risk with the best performing single-predictor model (early weight change at week 2) achieving a ROC-AUC score up to ∼ 0.85 (Lin et al., 2018). However, this result was limited by the small, highly controlled sample size and short follow-up duration, including only 67 Taiwanese participants that completed the six-week controlled trial, only 11 of whom gained significant weight. On another recent study including 1,234 children and adolescence aged 9-19 years treated with olanzapine, an overall ROC-AUC score of 0.74 was achieved in predicting significant weight gain but this performance was based on a pediatric cohort which also included non-olanzapine users as well, limiting both specificity to olanzapine and generalizability to adults (Lyu et al., 2024). Some of the existing studies also explore risk factors for weight gain in patients treated with olanzapine by analyzing data from different clinical trials (Fitzgerald, Sahm, Byrne, O’Connell, Ensor, Ní Dhubhlaing, et al., 2023; Huhn et al., 2019; Lipkovich et al., 2009). Key factors influencing weight increase in olanzapine treated patients across the literature were baseline BMI, where patients with higher baseline BMI tend to gain less weight and early weight increase being strongly predictive of long-term weight gain. While other factors such as younger age, female sex, non-smoking status, increased appetite early in treatment, and a diagnosis of schizophrenia spectrum disorder have been identified as potential contributing factors of increased weight gain (Eder et al., 2025; Harrison et al., 2017; Lipkovich et al., 2008, 2006; Kinon et al., 2001).

In our study we leveraged both statistical methods as well as machine learning approaches to explore and model BMI changes related to olanzapine in a Danish population psychiatric cohort of 6,254 individuals treated with olanzapine. We explored the feasibility and limitations of the current need in forecasting who is most at risk of significant weight gain after olanzapine prescription. We did this by using routinely collected clinical data from a psychiatric hospital setting. By integrating a wide range of available patient information (demographics, physical measurements, medications, diagnoses, lab tests and family history), we developed a predictive model of severe weight gain >5% of BMI increase, informing future efforts toward interventions for the prevention of patient weight gain on olanzapine which could also extend to other antipsychotics. Such information can offer clinicians an insight allowing them to tailor antipsychotic prescriptions based on individual profiles, so that healthcare providers can opt for closer monitoring, lifestyle changes, or alternative interventions, thus moving toward more personalized and proactive patient care.

## Results

### Elevated median BMI for individuals on olanzapine

We were interested in studying individuals treated with the antipsychotic medication olanzapine due to its previously reported weight gain inducing effects. Out of 153,663 adults in the psychiatric cohort, approximately 27,000 had been prescribed olanzapine. However, only 6,254 of these individuals (3,322 males and 2,932 females) had BMI measurements recorded while taking the medication and could be included in the study. This sub-cohort of 6,254 individuals formed the basis for our analysis (**Fig. 1**).

**Figure 1.**
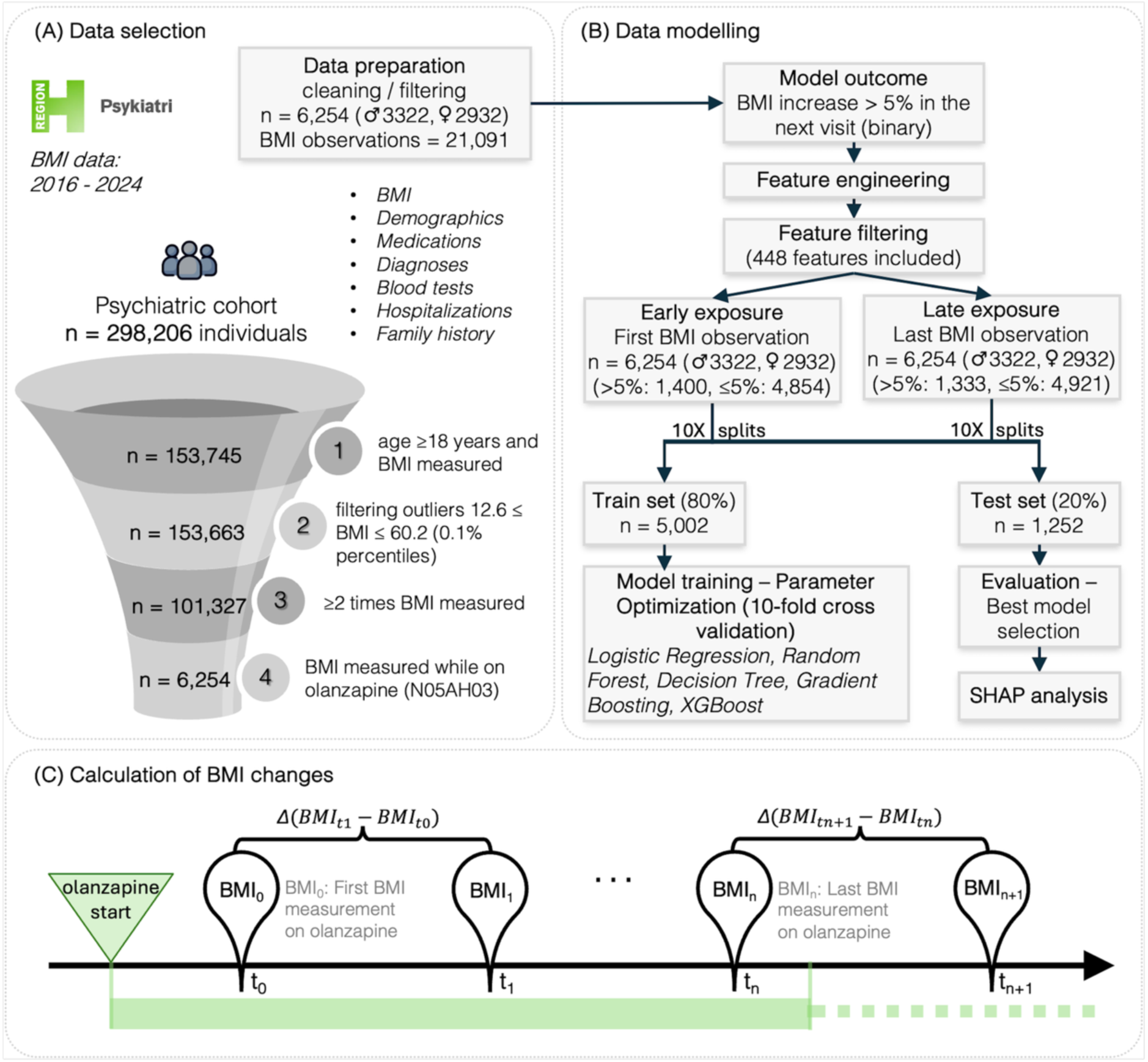
Overview of the study design, cohort selection, and machine learning pipeline. **(A)** Data selection process. Data were obtained from the population-based electronic health records covering the Capital Region of Denmark and the connected Region Zealand. After filtering for age >18 years, availability of BMI measurements, and olanzapine exposure a final cohort of 6,254 individuals was selected. **(B)** Machine Learning Modelling: Two distinct data subsets were created one for early exposure measurements and one for later exposure. Machine learning models were trained using the same modelling approach. The binary outcome was defined as a BMI increase >5% between two consecutive BMI measurements during olanzapine treatment. **(C)** Illustration of how the earliest and latest BMI measurements in relation to olanzapine treatment were defined. The predictive model outcome was to predict whether in the Δ(BMI) there is a >5% BMI increase from BMI_ti_ to BMI_ti+1_.

Compared to the entire adult psychiatric cohort where younger females are overrepresented (89,870 females and 63,793 males) (**Fig. 2A**) in the olanzapine sub-cohort the proportion of males and females was more balanced with ages between 18-94 years (**Fig. 2B**). The median BMI value in the entire cohort was 25.68 kg/m² for males and for females 24.8 kg/m² indicating that most male individuals were classified as overweight according to standard BMI thresholds, whereas females were generally within the upper limit of the normal weight category (**Fig. 2D**). The individuals on olanzapine had an elevated BMI and the median BMI for males was 26.54 kg/m^2^ and for females 26.04 kg/m^2^ (**Fig. 2E**), which was approximately one BMI point higher compared to individuals that never received olanzapine with the difference being statistically significant (Mann–Whitney U test, two-sided, p = 1.14*10^-26^) (**Fig. 2F**). The trajectory of BMI across the adult lifespan (**Fig. 2C**) revealed the broad patterns of weight change over time, with a general rise and then a declining trend after the age of 50 that did not differ from the pattern observed in the olanzapine sub-cohort. Even though the BMI trend did not differ, younger individuals treated with olanzapine exhibited higher BMI until middle age, which converged with that of those not treated with olanzapine after. Notably, the BMI variance which was calculated from each patient’s history of BMI measurements was also higher for the olanzapine group (**Supplementary Table 1**). In summary, individuals on olanzapine exhibited higher BMI especially until middle age and greater variability in BMI over time compared to the rest of the individuals in the cohort.

**Figure 2.**
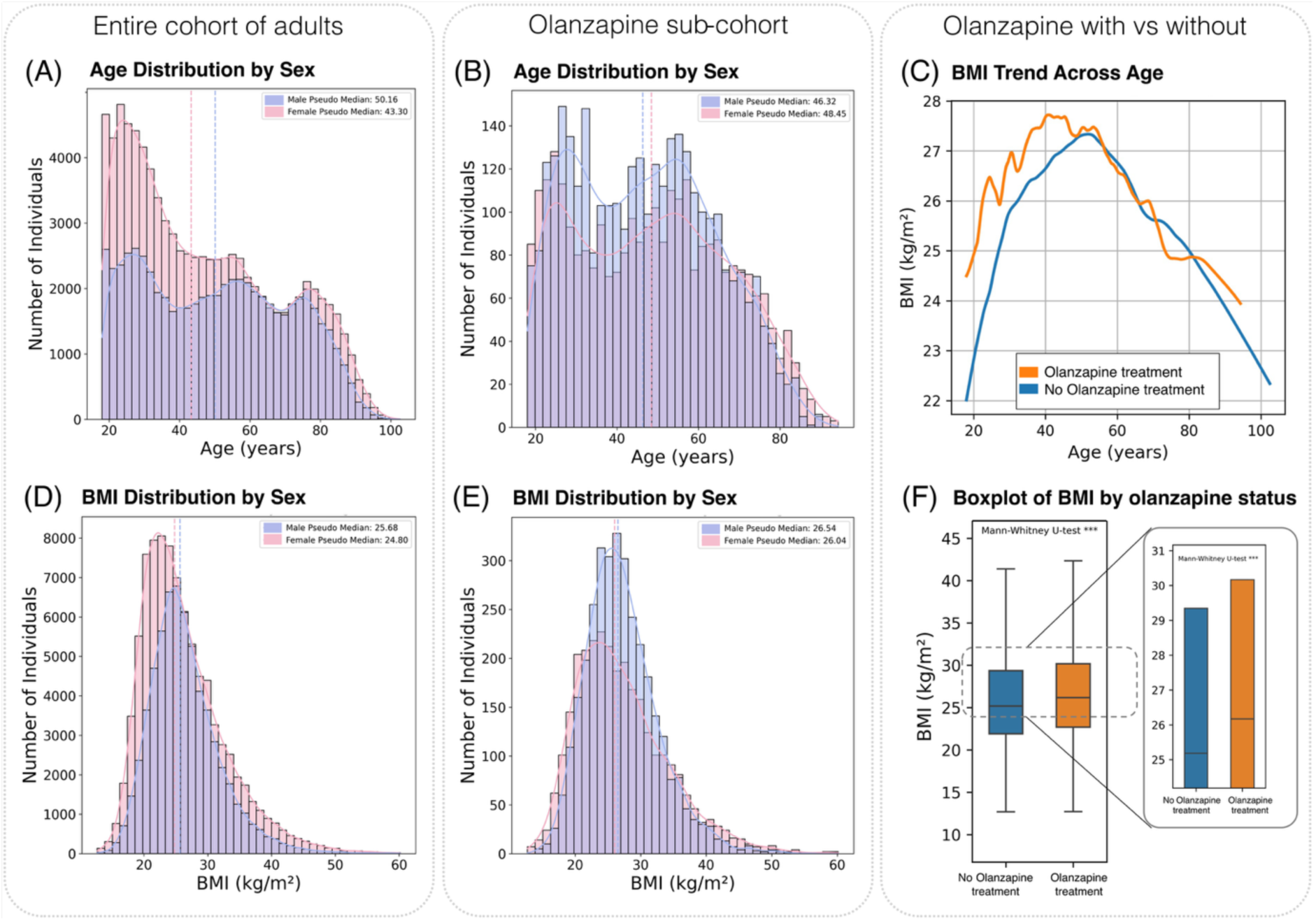
Exploratory plots of BMI, age and sex in the cohort. (**A, B)** Age distribution by sex in the entire cohort and the olanzapine sub-cohort respectively. Males are represented in purple and females in pink, with dashed lines in the corresponding colors indicating the median age values. **(C)** Trend of BMI across age in the cohort. The curves represent the LOWESS (locally weighted scatterplot smoothing) providing a smooth estimate of the overall trend, revealing an increase in BMI from early adulthood which is higher for young individuals treated with olanzapine, reaching a peak around middle age, and subsequently exhibiting a downward trend in later life. **(D, E)** BMI distribution by sex in both the overall cohort and the olanzapine sub-cohort. Males are represented in purple and females in pink, with dashed lines in the corresponding colors indicating the median BMI values. **(F)** Boxplot of BMI distributions comparing BMI of individuals on olanzapine and those that never received the medication. Each bar represents the range from the median BMI (horizontal line) to the 75th percentile, illustrating the upper half of the interquartile range.

### The rate of BMI change was higher for individuals on olanzapine

To investigate how the BMI changed for individuals treated with olanzapine, we analyzed the BMI measurements recorded during clinic visits focusing on the first visit after they initiated the medication and the last visit available while they are still being treated. At the first visit, individuals on olanzapine had a significantly elevated rate of BMI change compared to individuals not receiving the medication (on average +0.09 kg/m² per month) (Mann-Whitney U test, two-sided: p = 2.37×10⁻⁹), indicating a shift toward higher BMI gain over time in the olanzapine treated group (**Supplementary Table 1**). At the latest recorded BMI measurement, however, the rate of BMI change was not significantly different, indicating that the rate of BMI change stabilized over time. Overall, these findings confirm that individuals on olanzapine have a significantly elevated rate of BMI change in earlier observations compared to those who did not receive this medication, while at the latest recorded time point, the rate of BMI change appears to be converged to that of the rest of the cohort.

### BMI increase correlated positively with younger age and higher baseline BMI

To identify individuals at risk of experiencing a significant BMI increase, defined as a BMI increase of more than 5% between two consecutive BMI assessment visits, we first explored the correlations between variables in the dataset and our target outcome to better understand their potential predictive value. Spearman correlation analysis revealed a negative association between baseline BMI (Spearman ρ = -0.11) and age (Spearman ρ = -0.09) with future BMI increase, while several features as well as engineered ones, including time between BMI measurements (Spearman ρ = 0.22), total hospitalizations (Spearman ρ = 0.08), dosage (Spearman ρ = 0.07), duration of treatment (Spearman ρ = 0.07), BMI variance (Spearman ρ = 0.05) and anxiolytics exposure (Spearman ρ = 0.05), showed positive correlations with BMI increase >5% at the next visit (**Fig. 3**). Since many of the feature engineered variables are correlated with BMI increase >5% these findings highlight the importance of feature engineering to capture historical and temporal patterns relevant to the outcome.

**Figure 3.**
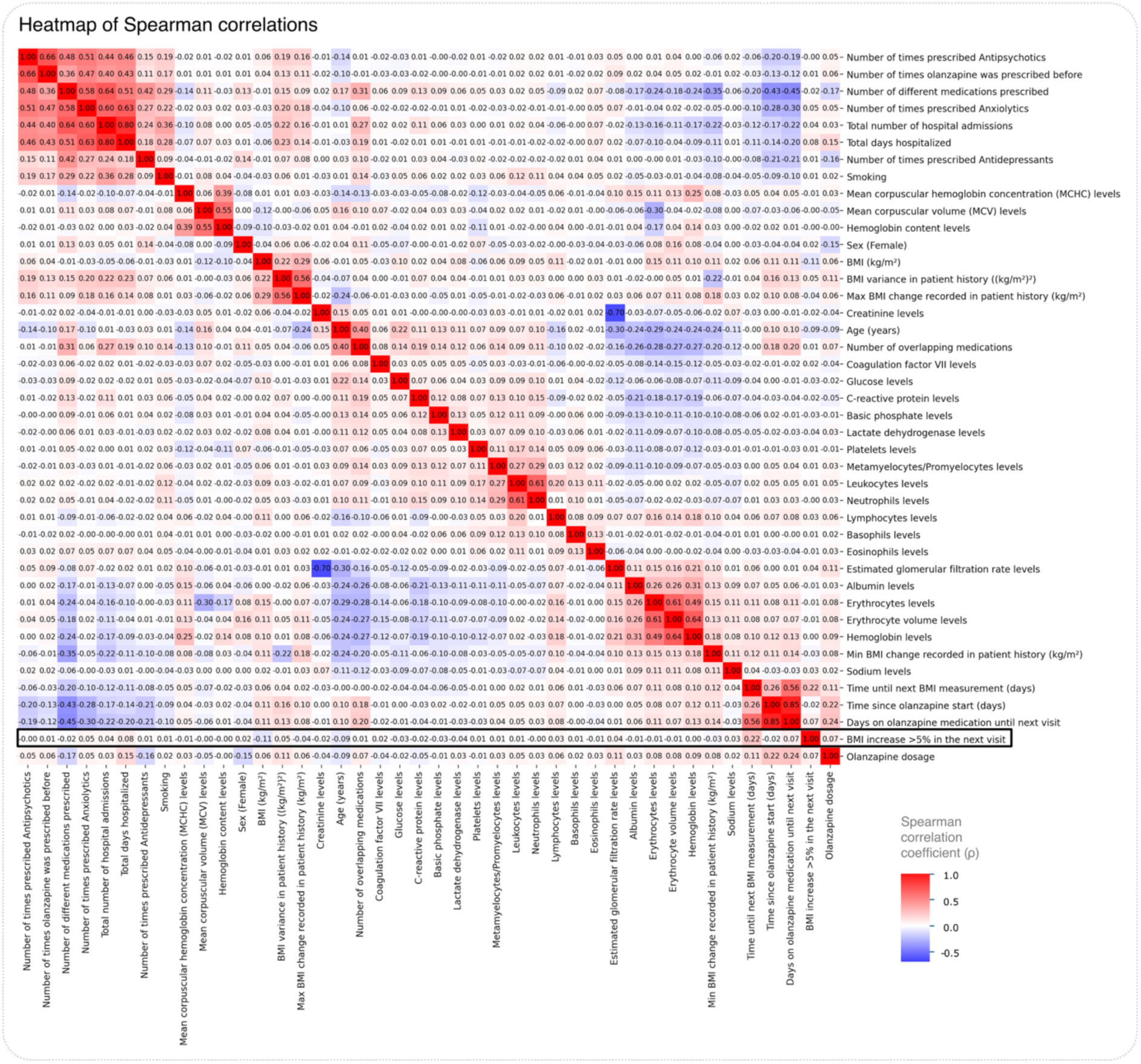
Correlation heatmap of engineered features and lab test variables. The heatmap illustrates pairwise Spearman correlation coefficients among variables analyzed in the olanzapine sub-cohort including engineered features, laboratory tests and medication-related information. Correlation coefficients are annotated within each cell, with stronger associations (positive: red color or negative: blue color) indicated by more intense colors. This analysis identifies interrelationships among potential predictors as well as predictors and the target variable outcome highlighting factors aiding interpretation and guiding the analysis.

### Olanzapine dosage correlated with male sex and younger age

Interestingly, when examining relationships among predictor variables olanzapine dosage showed stronger negative correlation with female sex (Spearman ρ = -0.15) and positive correlations with duration of the medication (Spearman ρ = 0.24), total number of days hospitalized (Spearman ρ = 0.15), and number of currently overlapping medications (Spearman ρ = 0.07), indicating that higher dosages, often part of more intensive treatment, relates to higher weight gain. Additionally, known relationships among predictors, such as abnormally higher glucose levels with older age (Spearman ρ = 0.22) and higher BMI (Spearman ρ = 0.10) emerged from the data. Finally, distinct positive and negative correlations among abnormal laboratory tests were observed. Particularly, liver enzymes such as lactate dehydrogenase and alkaline phosphatase (Spearman ρ = 0.13) demonstrated positive correlation, suggesting coordinated elevations which has been reported in different diseases (Grytczuk et al., 2020; Su et al., 2023), while kidney function markers creatinine and estimated glomerular filtration rate (eGFR) (Spearman ρ = -0.70) displayed a strong negative correlation, reflecting expected physiological relationships in kidney function where higher creatinine is associated with lower eGFR (Gansevoort et al., 2011; Xu et al., 2024). Notably, we evaluated the relationships between sex and various hematologic parameters both using the raw continuous laboratory values and after transforming those values into ordinal categories (1 = abnormal low, 2 = normal, 3 = abnormal high) allowing us to focus on deviations from the norm rather than absolute values. Analysis of the continuous measurements (**Supplementary Fig. 1**) confirmed well-established physiological differences between sexes; females typically exhibited lower erythrocyte counts (Spearman ρ = -0.27) and hemoglobin levels (Spearman ρ = -0.41), whereas they tended to have a higher transferrin values (Spearman ρ = 0.15) and mean corpuscular volume (MCV) values (Spearman ρ = 0.05) (Murphy, 2014). The correlation plot (**Fig. 3**) showed that females in the cohort we more likely to have ‘normal’ or ‘abnormally high’ levels of the blood parameters hemoglobin (Spearman ρ = 0.08), erythrocyte count (ρ = 0.08), and erythrocyte volume (ρ = 0.16), whereas males were more likely to have ‘normal’ or ‘abnormally low’ levels. These patterns in hematological tests are consistent with previous reports showing that antipsychotic treatment can influence these blood measurements (Jarari et al., 2025). Taken together these findings suggest that our dataset captures biologically and clinically meaningful associations, supporting its utility for predictive modeling.

### Predicting >5% BMI increase in the next visit after olanzapine early exposure

Recognizing the importance of identifying clinically significant BMI changes, we focused our approach to specifically predict more than 5% increase in BMI after olanzapine exposure. Throughout the machine learning modelling process, we trained different models: logistic regression, tree-based and ensemble models and found that XGBoost was the best performing model achieving a AUROC of 0.71 (F1 score positive class 0.43) (**Supplementary Table 2**), surpassing logistic regression (AUROC = 0.67) and other evaluated classifiers. The SHAP (SHapley Additive exPlanations) analysis highlighted several key predictive features including the ‘time until next BMI measurement’ (mean abs SHAP = 0.08), baseline ‘BMI’ (mean abs SHAP = 0.03), patient ‘age’ (mean abs SHAP = 0.03), cumulative ‘total number of days hospitalized’ (mean abs SHAP = 0.03), and ‘time since olanzapine start’ (mean abs SHAP = 0.02) (Lundberg & Lee, 2017). Specifically, prolonged intervals between follow-up visits, higher baseline BMI, and younger age were indicators of a BMI increase >5% (**Fig. 4A, Supplementary Table 3**). Additionally, features such as ‘olanzapine dosage’ (mean abs SHAP = 0.02) and the ‘number of overlapping medications’ (mean abs SHAP = 0.01) also showed contributions, emphasizing on the impact of medication regimens on BMI dynamics. Subgroup analyses demonstrated that model performance was consistent across sexes (female AUROC = 0.70, male AUROC = 0.71), while performance varied across age groups, with individuals in the group of 18-29 years showing better predictive accuracy (AUROC = 0.71) compared to the older group (AUROC = 0.68). These findings underscore the importance of clinical and demographic factors in predicting clinically relevant BMI changes, but it could also reflect the limitation that there was a smaller number of older individuals for the model to learn from (**Fig. 2B**).

**Figure 4.**
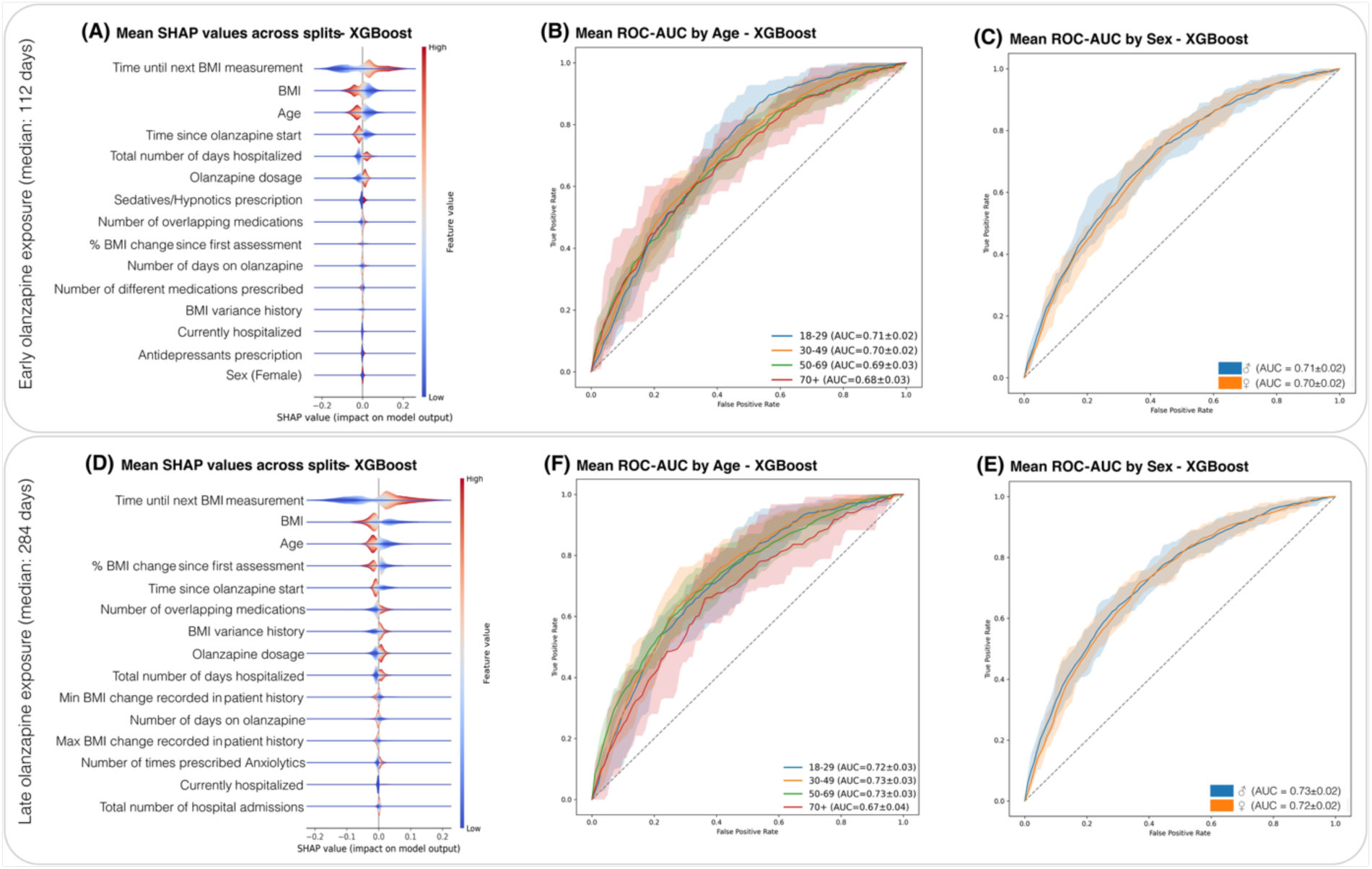
Machine learning modelling to predict >5% BMI increase after olanzapine exposure. Summary of the model’s predictive performance and influential features for early and late BMI assessment to the follow-up and BMI increase (>5%). **(A, D)** SHAP values (mean across 10 splits) for the most impactful features and best performing XGBoost model. Key factors driving the model predictions, notably ‘time until next BMI measurement’, ‘BMI’, ‘age’ and the ‘time since olanzapine start’ were the most informative features. **(B, F)** Comparison of ROC curves across different age groups (confidence intervals across 10 different splits) for the best performing XGBoost model. **(C, E)** Comparison of ROC curves across sexes (confidence intervals across 10 different splits) for the best performing XGBoost model, revealing consistent predictive accuracy across sexes.

Furthermore, we trained a regression model to forecast the exact magnitude of BMI changes; however, this approach did not produce reliable predictions and had low R-squared of ∼ 0.05 and a high RMSE of >3.5 kg/m² (**Supplementary Fig. 2**). Additionally, a binary classification model was also trained to predict weight gain or loss, but the outcome did not adequately capture the small weight increase versus decrease fluctuations (AUROC ∼ 0.56) (**Supplementary Fig. 3**). These results indicated that predicting a >5% BMI increase provided the most clinically meaningful and robust outcome compared to training our model to predict the magnitude and direction of all BMI changes that might not be clinically significant.

### Comparing predictions of BMI >5% increase after olanzapine late exposure

We trained a second model using the latest observations of BMI for each patient treated with olanzapine. Like above, the XGBoost model outperformed the other classifiers, achieving an overall AUROC of 0.72 (F1 score positive class 0.44) (**Supplementary Table 2**). SHAP analysis revealed that the most influential predictors were the ‘time until next BMI measurement’ (mean abs SHAP = 0.07), ‘BMI’ (mean abs SHAP = 0.03), ‘age’ (mean abs SHAP = 0.02), ‘% BMI change since the first assessment’ (mean abs SHAP = 0.02) and the ‘time since olanzapine start’ (mean abs SHAP = 0.01) (**Fig. 4D, Supplementary Table 3**). Furthermore, ROC curve stratifications by age and sex indicated subtle but noteworthy demographic effects: the model achieved the highest predictive performance in younger age groups (AUROC = 0.73), compared to the group of individuals ageing 70+ years (AUROC = 0.67) but still performed similarly in males (AUROC = 0.73) and females (AUROC = 0.72) (**Fig. 4E-F**).

Comparing these results to early olanzapine predictions above, we found that model performance were similar but key differences emerged in the order of feature importance and the specific features that ranked highest. In the late exposure model (**Fig. 4D**), ‘% BMI change since first assessment’ and ‘BMI variance history’ become more prominent features, reflecting the increasing relevance of historical BMI dynamics as more longitudinal data accumulate. ‘Min BMI change recorded in patient history’ and ‘Max BMI change recorded in patient history’ also appeared in the top 15 features (**Fig. 4D**), further supporting this shift towards historical weight trajectory being more informative in the late exposure model. In contrast, for the early prediction model (**Fig. 4A**), the model identified as key features acute clinical parameters and prescriptions, such as concurrent ‘Sedatives/Hypnotics prescription’ and other ‘Antidepressants prescription’. Overall, both analyses agreed on key predictors, however the late model incorporated more historical features, while the early model relied on baseline and immediate clinical information.

### A model with age, sex, BMI and time until next BMI measurement achieved similar performance

We also examined whether a model with a minimal set of features including ‘age’, ‘sex’, ‘BMI’, ‘time until the next BMI measurement’ could achieve a similar prediction performance. We re-trained the models using only these four variables. The stripped-down model yielded a median ROC-AUC of 0.69 for both early and late olanzapine exposure BMI measurements (**Supplementary Table 4**), very close to the previous models which included all the 448 variables (early ROC-AUC ∼ 0.71±0.01, late ROC-AUC ∼ 0.72±0.02). These results indicate that most of the discriminatory signals in predicting BMI increase resides in a handful of easily captured features while the rest of the clinical features only marginally improved performance.

## Discussion

In this study, we developed machine learning models leveraging real-world EHR data to predict clinically significant BMI increases (>5%) in patients undergoing olanzapine treatment in the early (median 112 days from olanzapine start) and the later stage of medication (median 284 days from olanzapine start). Our best-performing models, using XGBoost, achieved better predictive scores in the late stages on olanzapine (AUROC ∼ 0.72±0.02), compared to early predictions (AUROC ∼ 0.71±0.01) that could be reflecting the fact that the model has learned from the historical BMI measurements. The model’s performance suggests that there is potential utility of such modelling approaches for individualized risk stratification in clinical practice. However, key challenges are to have more frequent measurements and patient information about lifestyle factors in the EHRs. Key predictors identified in our analysis, ‘time until next BMI measurement’, ‘BMI’, ‘age’ and ‘olanzapine dosage’, align with existing literature on factors influencing antipsychotic-induced weight gain (Fitzgerald, Sahm, Byrne, O’Connell, Ensor, Dhubhlaing, et al., 2023; Kinon et al., 2001). But some other factors such as hospitalization frequency and length, use of sedatives, and prescriptions of anxiolytic medications have not been established before as risk factors.

We also asked whether a simpler model with less variables could achieve comparable performance. Using only age, sex, BMI, and the time until next BMI measurement, the XGBoost algorithm yielded an AUROC close to the full feature set (AUROC ∼ 0.69±0.01). The comparable results suggest that adding finer-grained clinical variables yields some additional predictive power for this outcome.

The negative correlation between baseline BMI and subsequent BMI increases, is consistent with previous studies indicating that patients with higher initial BMI tend to have relatively less BMI increase on olanzapine (Eder et al., 2025). This inverse relationship suggests that metabolic responses to olanzapine might differ based on baseline adiposity levels in patients with higher baseline weight. Similarly, age exhibited a negative correlation with BMI increase, in agreement with prior findings where younger individuals, particularly adolescents and young adults, are known to be more susceptible to substantial weight gain during antipsychotic treatment (Lee et al., 2011; Gebhardt et al., 2009). However, it may also reflect limitations in data availability since older individuals may have received olanzapine in the past priorly to the introduction of the current EHR system in 2016, and thus, first time initialization of olanzapine use might be more important than the age.

On the same note, our longitudinal data indicated that individuals experienced a higher rate of BMI increase early in their treatment course, with a subsequent plateau or stabilization in later measurements. This observation is in line with previous findings (Lipkovich et al., 2008), reporting that initial weight changes are predictive of longer-term outcomes where they found that early weight gain in the first 2–4 weeks of olanzapine treatment was a strong predictor of later substantial weight gain at around 30 weeks and reporting that patients with minimal early gain (e.g., <0.64 kg/m² BMI increase by week 3) had an 84% chance of avoiding large BMI increases in the next 7 months. That is reflected in the increased feature importance of BMI variance history in the late exposure model indicating that higher BMI variance was contributed to higher BMI increase.

The associations we observed between hospitalization frequency and increased olanzapine dosage suggest that more intensive psychiatric care and medication regimens coincide with greater BMI increases. This may reflect patients undergoing complex clinical trajectories characterized by multiple medication exposures, potentially amplifying metabolic dysregulation and weight gain risk (DE HERT et al., 2009; Nasrallah, 2008). Additionally, the increasing number of overlapping medications with olanzapine treatment (polypharmacy) also suggested greater risk for BMI increase. Our models highlighted that prior use of anxiolytics, and simultaneous use of sedatives/hypnotics and antidepressants also influence BMI increase, confirming hypotheses involving other psychotropics promoting weight gain (Correll et al., 2007; Kinon et al., 2001).

Our study demonstrates that using EHRs and modern machine learning algorithms, like XGBoost, can help predict which patients on olanzapine are most at risk for significant weight gain while at the same time highlighting potential risk factors (Hersh et al., 2013). Working with a large real-world cohort allowed us to build models that are translatable to clinical practice, but were affected by the common challenges with EHR data such as missing lifestyle information, irregular BMI recordings, and gaps in patient histories that constrained predictive performance and highlighted areas for improvement (Hripcsak et al., 2015). These findings emphasize the importance of standardized and consistent data collection by the clinicians, for ML models to be most useful in the clinic. Looking ahead, combining EHRs with other types of data, like lifestyle factors and omics data, will likely make it possible to identify high-risk patients more accurately and support more personalized care to help prevent weight-related complications.

## Methods

### Data Collection and Patient Selection

In this study, we leveraged electronic health record (EHR) data (de-identified) from psychiatric clinics in the Capital Region of Denmark and Region Zealand, spanning from May 2016 to January 2024. Our objective was to develop a predictive model capable of forecasting future BMI changes for individuals with psychiatric contacts. To ensure meaningful analysis of BMI changes, we included all individuals with at least two BMI assessments, as a minimum of two measurements was necessary to assess BMI changes over time while the patients were treated with olanzapine (**Fig. 1A**). Individuals under the age of 18 were excluded from the analysis to avoid the confounding effects of growth and development during childhood development, which involve unique trajectories in height, muscle mass, and fat distribution that is not directly comparable to adults (Rzehak et al., 2017). After applying these selection criteria 6,254 individuals were selected to be part of the study cohort.

### Feature engineering and preprocessing

To enhance the predictive performance with information about the history of each patient, several features that were derived from the EHR data were engineered to reflect the historical measurements of BMI, hospitalizations and medication exposure (**Supplementary Table 5**). Given the irregular intervals of BMI measurements, temporal features were engineered to capture historical time-dependent variations and avoid potential measurement biases. Temporal and BMI dynamics help account for time inconsistencies and past fluctuations, while hospitalization and medication exposure information provide additional layers of insight into clinical factors affecting the BMI increase. BMI changes were defined as the percentage difference between consecutive BMI measurements (ΔBMI (%) = (BMI_ti+1_ − BMI_ti_) / BMI_ti_ × 100), and the rate of change was calculated by dividing the BMI difference by the time that elapsed between the two measurements (ΔBMI (rate) = (BMI_ti+1_ − BMI_ti_) / (t_i+1_ − t_i_)). For each of the 6,254 patients we calculated the BMI change from their first available BMI measurement on olanzapine to the next (BMI_t0_ → BMI_t1_ early exposure - median olanzapine exposure 112 days) and from their last available BMI measurement on olanzapine to the next (BMI_tn_ → BMI_tn+1_ late exposure - median olanzapine exposure 284 days) (**Supplementary Fig. 4A, 4C**). The predictive model outcome was to predict whether in the ΔBMI (%) there was a >5% BMI increase from BMI_ti_ to BMI_ti+1._ The dataset underwent preprocessing to handle missing values and outliers (**Supplementary Methods**). Missing values were imputed using either the median or the most frequent value, depending on the variable’s distribution. Features with more than 50% missing values across the samples were excluded from the analysis; the only exception was the set of historical BMI engineered features (59 % missing at the first olanzapine assessment), which were retained to explore information on historical weight trajectories that were considered clinically relevant.

### Feature Extraction, Selection and Modeling Approach

A set of 448 features was extracted from the EHR data, including demographic variables (e.g., age, sex), feature engineered historical BMI metrics (e.g., variance, percentage change, max/min deviations), medication exposure (e.g., days on medication, number of antipsychotic prescriptions), and clinical history (e.g., hospitalization counts and durations) (**Supplementary Table 6, Supplementary Table 7**). Categorical variables (medication and diagnoses codes) were one hot encoded so that each medication or disease code corresponds to a separate binary feature column, while continuous variables were z-standardized to facilitate model convergence (Pedregosa et al., 2018). Based on preliminary analysis and preprocessing we removed features with more than 50% missingness and kept only the top 200 most frequent medication (ATC) and diagnosis (SKS) codes to avoid introducing sparsity in our table. Initially, we applied a continuous regression approach to predict the BMI change in follow-up, which yielded unsatisfactory results (R-squared of 0.05 and RMSE of 3.5 kg/m²) (**Supplementary Fig. 2**). We then explored a binary classification model to predict weight gain versus loss/no change, but it failed to capture subtle, BMI fluctuations (AUROC ∼ 0.56) (**Supplementary Fig. 3**). Consequently, we refined our approach to the model outcome as >5% BMI increase between consecutive clinic visits, thereby transforming the problem into a meaningful classification task since a 5% increase in BMI was recognized as a critical indicator of significant weight gain that may correlate with health risks.

### Machine Learning Model Development

Several machine learning algorithms were evaluated for a binary classification outcome prediction (Logistic Regression (Wright, 1995), Random Forest (Breiman, 2001), Decision Tree (Quinlan, 1990), Gradient Boosting (He et al., 2019) and XGBoost (Chen & Guestrin, 2016)), using the selected features as inputs. XGBoost emerged as the best performing model for modeling significant BMI increase using EHR tabular data. The dataset was split into a training set (80% - 5,002 samples) and an independent test set (20% - 1,252 samples); this outer split was repeated 10 times with different random seeds. Within each training set, hyper-parameters for all five algorithms were optimized via grid search and 5-fold stratified cross-validation (scored by ROC-AUC). The best estimator from each grid was re-trained on the full training data and evaluated on the held-out test set, which was kept entirely separate and reserved for final performance evaluation on each of the models. The performance metrics and SHAP values for the best model across the different algorithms were summarized by averaging the results across the ten splits (Lundberg & Lee, 2017). The data preprocessing and modelling were performed using Python version 3.13.2, scikit-learn version 1.6.1 and xgboost 3.0.0 libraries (Chen & Guestrin, 2016; Pedregosa et al., 2018; Python Development Team, 2025).

### Model Evaluation

We evaluated modelling performance using measures of discrimination, calibration, and overall predictive accuracy. Discrimination was primarily quantified by the area under the Receiver Operating Characteristic curve (ROC), which reflects the probability that the model correctly assigns a higher predicted probability of BMI increase to a patient who experiences this event relative to one who does not. In our context, an Area Under the Curve (AUC) of 0.5 would indicate performance no better than chance, whereas an AUC of 1.0 would indicate perfect discrimination (Bradley, 1997; Richardson et al., 2024). Alongside the ROC-AUC, we reported conventional metrics such as Accuracy (the overall proportion of correct predictions), Sensitivity (the model’s ability to correctly identify patients with significant BMI increases), Specificity (the model’s ability to correctly identify patients without a substantial BMI increase), Precision (the positive predictive value), and the F1-score (the harmonic mean of precision and recall – both weighted and macro). Furthermore, we conducted exploratory subgroup analyses to assess generalizability stratifying by sex and age. SHAP feature importance analysis was implemented to gain insights into the underlying factors that drove the model decisions. SHAP assigns each feature an importance value, its ‘Shapley value’, that quantifies how much each feature has contributed to the predictions (Lundberg & Lee, 2017). The results of the SHAP analysis on the test set were averaged across the 10 data splits and provided valuable insights into the underlying factors that drove the model decisions and are associated with these predictions.

## Data Availability and ethics

The project was approved by the Data Protection Agency of the Capital Region of Denmark and the Danish Society for Patient Safety (approval numbers: P-2020-101 and R-22002033), and all data processing was conducted in compliance with stringent data protection regulations to safeguard patient confidentiality. According to Danish regulations the analyses do not require informed consent. The data used in this study are not publicly available in order to comply with Danish data protection regulations and safeguard patient confidentiality. Access to the data may be granted to qualified researchers who meet the criteria for access to confidential health data, subject to approval by the relevant Danish authorities.

## Code Availability

The code for the study presented in this paper is openly accessible and can be found in its entirety on the GitHub repository at https://github.com/fotiniak/bmipred. The code was tailored specifically to the data structure available for our system, and it might therefore not be easily generalizable to other data sources.

## Acknowledgements

We would like to thank the group members Jonas Meisner, Enric Cristobal Coppulo and Terne Sasha Thorn Jakobsen for their feedback and fruitful discussions. We would also like to thank the Novo Nordisk Foundation Center for Basic Metabolic Research and the Copenhagen Research Centre for Biological and Precision Psychiatry for their support.

## Author contribution

M.E.B. and S.R. conceptualized and initiated the study. F.A, S.R. and M.E.B. guided the analysis. F.A. wrote the code and performed the analysis. F.A. wrote the manuscript with contributions from all coauthors. All authors read and approved the final version of the manuscript.

## Supplementary information

Additional file 1. Supplementary Information.docx

Additional file 2. Supplementary Table 6.xlsx

Additional file 3. Supplementary Table 7.xlsx

