## Supplementary Information for "Predicting Olanzapine Induced BMI increase using Machine Learning on population-based Electronic Health Records"

**Supplementary Table 1: Mann-Whitney U test results.** Comparison of individuals treated with olanzapine vs those without olanzapine treatment across two data subsets: The first recorded BMI measurement per individual (representing early olanzapine exposure) and the last recorded BMI measurement per individual (representing late olanzapine exposure). At the early olanzapine exposure group both BMI and the rate of BMI change were significantly higher in the olanzapine group (p < 0.001) and in the later olanzapine exposure BMI and BMI variance were significantly higher in the olanzapine group (p < 0.001), but the rate of BMI change did not differ significantly (p = 0.805).

| Variable | No olanzapine treatment | | Olanzapine treatment | | p-value |
| --- | --- | --- | --- | --- | --- |
|  | N | Mean | N | Mean |  |
| BMI (kg/m²) | 86529 | 26.352 | 6254 | 26.957 | 3.05 × 10⁻²¹ (<0.001) |
| rate of BMI change (kg/m² per month) | 86529 | 0.041 | 6254 | 0.092 | 2.37 × 10⁻⁹ (<0.001) |
| Variable | No olanzapine treatment | | Olanzapine treatment | | p-value |
|  | N | Mean | N | Mean |  |
| BMI (kg/m²) | 86529 | 26.493 | 6254 | 27.212 | 1.14 × 10⁻²⁶ (<0.001) |
| rate of BMI change (kg/m² per month) | 86529 | 0.017 | 6254 | 0.008 | 8.05 × 10⁻¹  (not significant) |
| BMI variance ((kg/m²)²) | 59458 | 2.938 | 4777 | 3.628 | 9.68 × 10⁻⁵⁴ (<0.001) |

**Supplementary Table 2: Performance metrics of five machine-learning classifiers in predicting a binary outcome (>5% BMI increase) using 448 features.** Models tested include Logistic Regression, Random Forest, Decision Tree, Gradient Boosting, and XGBoost. Reported metrics include AUROC (area under the ROC curve), F1 Score (positive class), F1 Score (weighted), Accuracy, Sensitivity (true positive rate), Specificity (true negative rate), PPV (positive predictive value), NPV (negative predictive value), and MCC (Matthews correlation coefficient) and the 95% CI across the 10 data splits.

| **First BMI measurement on olanzapine** | | | | | | | | | |
| --- | --- | --- | --- | --- | --- | --- | --- | --- | --- |
| **Model** | **AUROC** | **F1 (positive)** | **F1 (weighted)** | **Accuracy** | **Sensitivity** | **Specificity** | **PPV** | **NPV** | **MCC** |
| Decision Tree | 0.621 (0.589–0.642) | 0.357 (0.302–0.4) | 0.671 (0.638–0.698) | 0.653 (0.607–0.69) | 0.436 (0.305–0.58) | 0.716 (0.614–0.8) | 0.308 (0.287–0.341) | 0.816 (0.798–0.836) | 0.137 (0.096–0.185) |
| Gradient Boosting | 0.7 (0.683–0.717) | 0.412 (0.382–0.44) | 0.728 (0.705–0.746) | 0.724 (0.687–0.748) | 0.433 (0.36–0.533) | 0.808 (0.731–0.857) | 0.398 (0.364–0.433) | 0.832 (0.821–0.845) | 0.235 (0.207–0.269) |
| Logistic Regression | 0.673 (0.652–0.694) | 0.423 (0.401–0.451) | 0.647 (0.589–0.679) | 0.617 (0.553–0.653) | 0.626 (0.54–0.691) | 0.614 (0.515–0.681) | 0.321 (0.29–0.346) | 0.851 (0.835–0.867) | 0.203 (0.166–0.247) |
| Random Forest | 0.697 (0.677–0.716) | 0.419 (0.39–0.457) | 0.717 (0.702–0.742) | 0.707 (0.683–0.749) | 0.476 (0.367–0.569) | 0.773 (0.724–0.859) | 0.38 (0.356–0.433) | 0.837 (0.824–0.855) | 0.232 (0.198–0.271) |
| **XGBoost** | **0.705 (0.684–0.724)** | **0.429 (0.374–0.471)** | **0.723 (0.699–0.751)** | **0.714 (0.681–0.752)** | **0.482 (0.347–0.581)** | **0.781 (0.728–0.866)** | **0.392 (0.35–0.443)** | **0.84 (0.819–0.86)** | **0.247 (0.193–0.303)** |

| **Latest BMI measurement on olanzapine** | | | | | | | | | |
| --- | --- | --- | --- | --- | --- | --- | --- | --- | --- |
| **Model** | **AUROC** | **F1 (positive)** | **F1 (weighted)** | **Accuracy** | **Sensitivity** | **Specificity** | **PPV** | **NPV** | **MCC** |
| Decision Tree | 0.638 (0.611–0.668) | 0.378 (0.339–0.405) | 0.676 (0.624–0.716) | 0.652 (0.586–0.704) | 0.499 (0.376–0.618) | 0.694 (0.585–0.785) | 0.31 (0.279–0.349) | 0.837 (0.82–0.853) | 0.168 (0.132–0.213) |
| Gradient Boosting | 0.717 (0.694–0.75) | 0.435 (0.389–0.491) | 0.746 (0.722–0.777) | 0.739 (0.712–0.774) | 0.471 (0.405–0.524) | 0.812 (0.781–0.847) | 0.405 (0.358–0.474) | 0.85 (0.835–0.864) | 0.268 (0.207–0.346) |
| Logistic Regression | 0.693 (0.662–0.734) | 0.437 (0.411–0.484) | 0.705 (0.686–0.734) | 0.682 (0.658–0.717) | 0.58 (0.502–0.647) | 0.71 (0.662–0.768) | 0.352 (0.328–0.391) | 0.862 (0.85–0.882) | 0.249 (0.209–0.315) |
| Random Forest | 0.715 (0.685–0.745) | 0.428 (0.369–0.49) | 0.748 (0.737–0.757) | 0.745 (0.718–0.759) | 0.458 (0.335–0.614) | 0.823 (0.746–0.871) | 0.413 (0.394–0.43) | 0.85 (0.828–0.878) | 0.27 (0.221–0.33) |
| **XGBoost** | **0.725 (0.704–0.752)** | **0.442 (0.395–0.486)** | **0.743 (0.726–0.756)** | **0.734 (0.708–0.752)** | **0.498 (0.407–0.622)** | **0.798 (0.733–0.842)** | **0.402 (0.372–0.425)** | **0.855 (0.837–0.877)** | **0.275 (0.219–0.323)** |

**Supplementary Table 3. SHAP-derived feature importance for the models trained on the first BMI measurement while on olanzapine and latest respectively.** The table lists the **20 most influential predictors,** ranked by the **mean absolute SHAP value (|SHAP|)** calculated on the external test set. **Mean |SHAP|** reflects the **average magnitude** with which a feature moves the model’s log-odds of experiencing a >5 % BMI increase irrespective of direction. **Std |SHAP|** is the corresponding **standard deviation**, indicating how much that influence varies from one patient to another.

**Early BMI measurement table SHAP values**

| **Feature** | **Mean abs shap** | **Std abs shap** |
| --- | --- | --- |
| Time until next BMI measurement (days) | 0.07480597 | 0.051834354 |
| BMI (kg/m²) | 0.032773624 | 0.024882269 |
| Age (years) | 0.028844552 | 0.020350003 |
| Time since olanzapine start | 0.022621178 | 0.014485938 |
| Total days hospitalized | 0.020653047 | 0.01140049 |
| Olanzapine dosage | 0.016508175 | 0.010604324 |
| ATC_N05C (Sedatives/Hypnotics) | 0.007032993 | 0.004694486 |
| % BMI change since first measurement | 0.005587809 | 0.009143419 |
| Number of currently overlapping medications | 0.005542828 | 0.006115655 |
| Days on olanzapine medication until next visit | 0.00487173 | 0.006547601 |
| Number of different medications prescribed | 0.004697662 | 0.005045589 |
| BMI variance history ((kg/m²)²) | 0.004573662 | 0.006353311 |
| ATC_N06A (Antidepressants) | 0.003462844 | 0.002826937 |
| Currently hospitalized | 0.003419246 | 0.005432009 |
| Sex (Female) | 0.002700925 | 0.002337712 |
| Albumin levels | 0.002225935 | 0.003061115 |
| Number of times on anxiolytics prescription | 0.002082836 | 0.002500024 |
| Total number of hospital admissions | 0.002082356 | 0.002801263 |
| Min BMI change recorded in patient history (kg/m²) | 0.001847004 | 0.003186901 |
| Max BMI change recorded in patient history (kg/m²) | 0.001415701 | 0.003102294 |

**Last BMI measurement table SHAP values**

| **Feature** | **Mean abs shap** | **Std abs shap** |
| --- | --- | --- |
| Time until next BMI measurement (days) | 0.066786212 | 0.04365553 |
| BMI (kg/m²) | 0.033043597 | 0.023320041 |
| Age (years) | 0.01960858 | 0.014340721 |
| % BMI change since first measurement | 0.015685857 | 0.01185269 |
| Time since olanzapine start | 0.014227409 | 0.01058923 |
| Number of currently overlapping medications | 0.011595254 | 0.011727379 |
| BMI variance history ((kg/m²)²) | 0.011565635 | 0.01054931 |
| Total days hospitalized | 0.010030262 | 0.006851471 |
| Olanzapine dosage | 0.009951145 | 0.007146645 |
| Min BMI change recorded in patient history (kg/m²) | 0.007172647 | 0.006927741 |
| Max BMI change recorded in patient history (kg/m²) | 0.006913718 | 0.010846624 |
| Times on anxiolytics prescription | 0.005596561 | 0.004870501 |
| Number of days on olanzapine | 0.004453084 | 0.005101668 |
| Currently hospitalized | 0.004420431 | 0.006927088 |
| Glucose levels | 0.003177338 | 0.00349751 |
| Total number of hospital admissions | 0.002777939 | 0.003957158 |
| ATC_A02A (Antacids) | 0.002461199 | 0.003939902 |
| Number of different medications prescribed | 0.002198529 | 0.002235788 |
| ATC_N05C (Sedatives/Hypnotics) | 0.001905528 | 0.00216973 |
| SKS_DZ71 (Healthcare System Contact) | 0.0018549 | 0.003593318 |

**Supplementary Table 4: Performance metrics of five machine-learning classifiers in predicting a binary outcome (>5% BMI increase) using only ‘age’, ‘sex’, ‘BMI’ and ‘time until the next BMI measurement’ as features.** Models tested include Logistic Regression, Random Forest, Decision Tree, Gradient Boosting, and XGBoost. Reported metrics include AUROC (area under the ROC curve), F1 Score (positive class), F1 Score (weighted), Accuracy, Sensitivity (true positive rate), Specificity (true negative rate), PPV (positive predictive value), NPV (negative predictive value), and MCC (Matthews correlation coefficient) and the 95% CI across the 10 data splits.

| **First BMI measurement on olanzapine** | | | | | | | | | |
| --- | --- | --- | --- | --- | --- | --- | --- | --- | --- |
| **Model** | **AUROC** | **F1 (positive)** | **F1 (weighted)** | **Accuracy** | **Sensitivity** | **Specificity** | **PPV** | **NPV** | **MCC** |
| Decision Tree | 0.662 (0.64–0.688) | 0.424 (0.412–0.458) | 0.675 (0.665–0.698) | 0.65 (0.635–0.679) | 0.577 (0.523–0.689) | 0.671 (0.62–0.724) | 0.336 (0.325–0.357) | 0.847 (0.839–0.874) | 0.213 (0.193–0.26) |
| Gradient Boosting | 0.688 (0.659–0.712) | 0.441 (0.414–0.466) | 0.673 (0.635–0.698) | 0.646 (0.602–0.675) | 0.624 (0.557–0.699) | 0.652 (0.574–0.7) | 0.342 (0.318–0.363) | 0.858 (0.84–0.875) | 0.235 (0.197–0.272) |
| Logistic Regression | 0.665 (0.644–0.692) | 0.425 (0.408–0.442) | 0.607 (0.588–0.62) | 0.572 (0.553–0.586) | 0.705 (0.655–0.736) | 0.534 (0.504–0.561) | 0.304 (0.294–0.316) | 0.863 (0.848–0.876) | 0.2 (0.171–0.231) |
| Random Forest | 0.689 (0.662–0.714) | 0.439 (0.411–0.463) | 0.67 (0.642–0.693) | 0.642 (0.61–0.667) | 0.626 (0.537–0.693) | 0.647 (0.588–0.693) | 0.339 (0.318–0.362) | 0.858 (0.838–0.869) | 0.232 (0.194–0.269) |
| XGBoost | 0.692 (0.663–0.717) | 0.441 (0.413–0.472) | 0.645 (0.614–0.68) | 0.614 (0.58–0.654) | 0.683 (0.558–0.774) | 0.594 (0.529–0.682) | 0.327 (0.31–0.344) | 0.868 (0.84–0.896) | 0.233 (0.193–0.283) |

| **Last BMI measurement on olanzapine** | | | | | | | | | |
| --- | --- | --- | --- | --- | --- | --- | --- | --- | --- |
| **Model** | **AUROC** | **F1 (positive)** | **F1 (weighted)** | **Accuracy** | **Sensitivity** | **Specificity** | **PPV** | **NPV** | **MCC** |
| Decision Tree | 0.651 (0.604–0.69) | 0.393 (0.356–0.455) | 0.708 (0.664–0.743) | 0.693 (0.636–0.731) | 0.467 (0.387–0.535) | 0.754 (0.671–0.811) | 0.342 (0.295–0.4) | 0.839 (0.826–0.86) | 0.2 (0.15–0.285) |
| Gradient Boosting | 0.691 (0.672–0.711) | 0.427 (0.399–0.463) | 0.715 (0.701–0.736) | 0.696 (0.678–0.719) | 0.532 (0.469–0.611) | 0.741 (0.702–0.768) | 0.358 (0.338–0.391) | 0.854 (0.841–0.871) | 0.24 (0.205–0.29) |
| Logistic Regression | 0.663 (0.633–0.692) | 0.411 (0.388–0.436) | 0.664 (0.639–0.682) | 0.633 (0.604–0.654) | 0.601 (0.545–0.657) | 0.641 (0.601–0.677) | 0.313 (0.293–0.33) | 0.856 (0.843–0.872) | 0.202 (0.165–0.24) |
| Random Forest | 0.692 (0.674–0.711) | 0.43 (0.409–0.459) | 0.709 (0.688–0.72) | 0.688 (0.66–0.705) | 0.552 (0.494–0.652) | 0.725 (0.665–0.762) | 0.353 (0.337–0.369) | 0.857 (0.845–0.876) | 0.241 (0.215–0.28) |
| XGBoost | 0.69 (0.672–0.712) | 0.426 (0.409–0.458) | 0.707 (0.689–0.731) | 0.686 (0.661–0.715) | 0.549 (0.474–0.652) | 0.723 (0.665–0.772) | 0.35 (0.328–0.382) | 0.856 (0.844–0.878) | 0.236 (0.208–0.282) |

**Supplementary Table 5. Table of engineered features.** Summary of the set of engineered features used for modeling and analysis, organized into four main categories: BMI History, Medication Exposure, Clinical History, and Temporal Dynamics. Each feature is listed alongside with a brief description of how it is defined or derived from the raw data. By capturing multiple dimensions—body weight historical trends, medication use, clinical background, and temporal factors—we can capture a comprehensive view of each patient’s longitudinal health profile. Such multidimensional data can enhance predictive modeling and help identify patterns associated with treatment responses or disease progression.

| **Category** | **Feature** | **Type** | **Description** |
| --- | --- | --- | --- |
| **BMI History** | BMI variance history | Numeric | Variability in previous BMI measurements. |
|  | % BMI change since first measurement | Numeric | Percentage change in BMI from the first recorded measurement in the clinic. |
|  | Max BMI change recorded in patient history (kg/m²) | Numeric | The highest deviation in BMI over historical records before. |
|  | Min BMI change recorded in patient history (kg/m²) | Numeric | The lowest deviation in BMI over historical records before. |
| **Medication Exposure** | Days on olanzapine medication until next visit | Numeric | Duration of olanzapine use. |
|  | Times olanzapine was prescribed before | Numeric | Count of previous olanzapine prescriptions. |
|  | Olanzapine status in the next assessment | Boolean | Indicator of whether the patient is still receiving olanzapine at the next follow-up. |
|  | Times on antipsychotics prescription | Numeric | Count of prescriptions for antipsychotic medications. |
|  | Times on antipsychotics prescription | Numeric | Count of prescriptions for anxiolytic medications. |
|  | Times on antidepressants prescription | Numeric | Count of prescriptions for antidepressant medications. |
|  | Number of currently overlapping medications | Numeric | Count of the different medications prescribed simultaneously. |
|  | Number of different medications prescribed | Numeric | Count of unique medications the patient has been exposed to over their medical history. |
| **Clinical History** | Total number of hospital admissions | Numeric | Total number of psychiatric hospitalizations. |
|  | Total days hospitalized | Numeric | Cumulative number of days spent in the hospital across hospitalizations. |
|  | Currently Hospitalized | Boolean | Indicator of whether the patient was hospitalized when the baseline BMI was measured. |
| **Temporal Dynamics** | Days on olanzapine medication until next visit | Numeric | Time elapsed from the start of olanzapine treatment until baseline BMI measurement. |
|  | Time until next BMI measurement (days) | Numeric | Time elapsed between current and next BMI measurements. |

**Supplementary Table 6. List of all features in the early olanzapine exposure table**

Supplementary Table 6.xlsx

**Supplementary Table 7. List of all features in the late olanzapine exposure table.**

Supplementary Table 7.xlsx

**Supplementary Figure 1: Correlation of Sex (Female) with Laboratory Test Results in the olanzapine sub-cohort.** Pearson correlation coefficients between sex (female) and a variety of laboratory test results, including hematological and biochemical markers. Each bar represents the correlation coefficient between female sex and a specific laboratory test. The colors represent the strength of the correlation, where blue corresponds to negative correlations and red to positive correlation. Some of the strongest positive correlations are observed with tests like platelets and transferrin while a negative correlation is seen with tests such as uric acid and hemoglobin, where higher levels are associated with lower likelihood of being female.

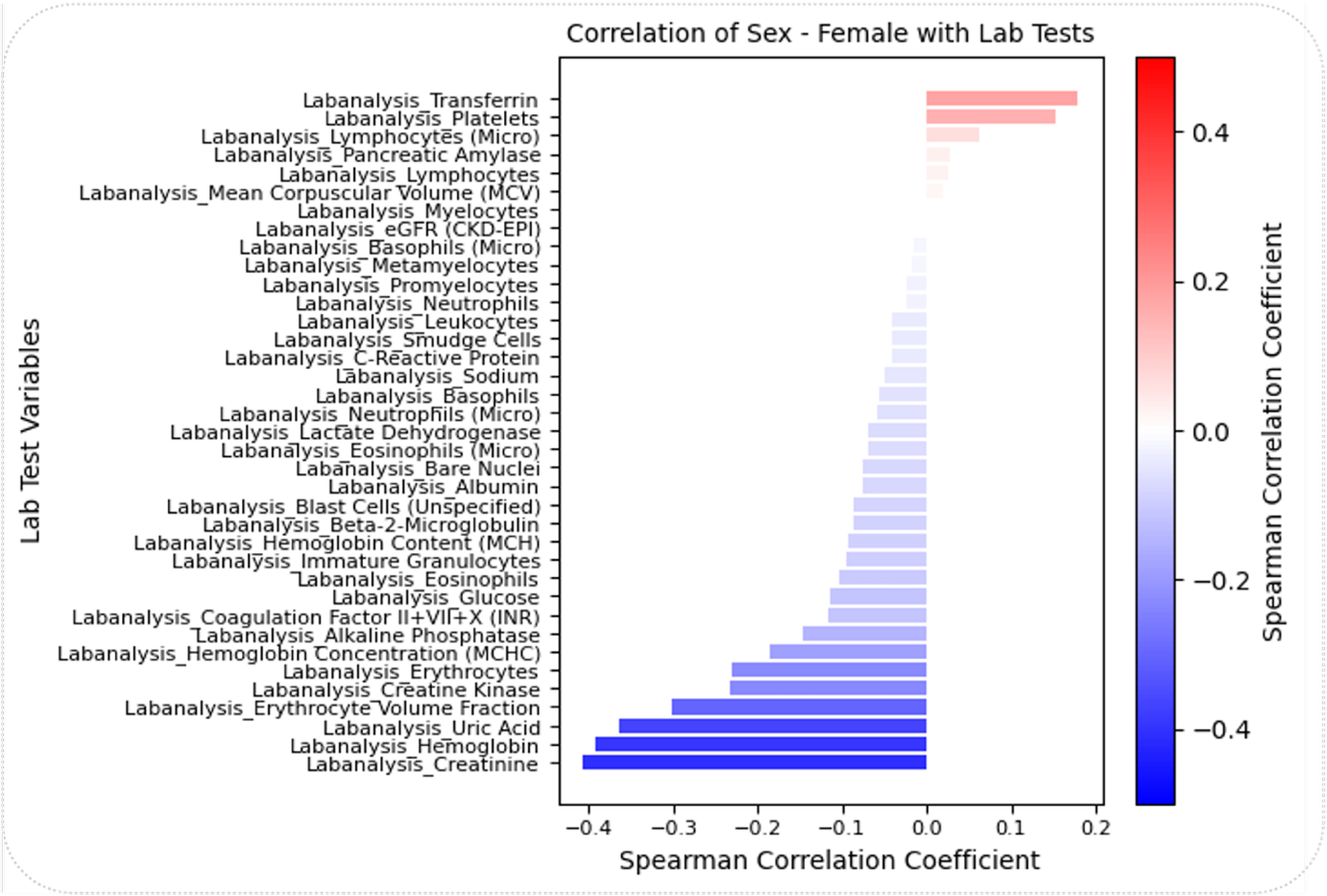

**Supplementary Figure 2: Machine Learning model outcome (R^2^) for the prediction of the numerical BMI change values in the next visit for each patient after the first BMI assessment while treated with olanzapine.** Comparison of regression model’s predicted values (y-axis) versus the true (actual) values (x-axis). Each point corresponds to a single prediction of the BMI change continuous value, and colors denote different models (LinearRegression, Ridge, Lasso, KNNRegressor, DecisionTreeRegressor, GradientBoostingRegressor, MLPRegressor, XGBoostRegressor). The dashed diagonal line indicates perfect agreement (predicted = actual). R² values in parentheses highlight each model’s fit; points clustering closer to the diagonal reflect higher predictive accuracy. Notably, the GradientBoostingRegressor (R² ≈ 0.05) is the best performing, while other methods exhibit lower or even negative R² scores, indicating poorer fits.

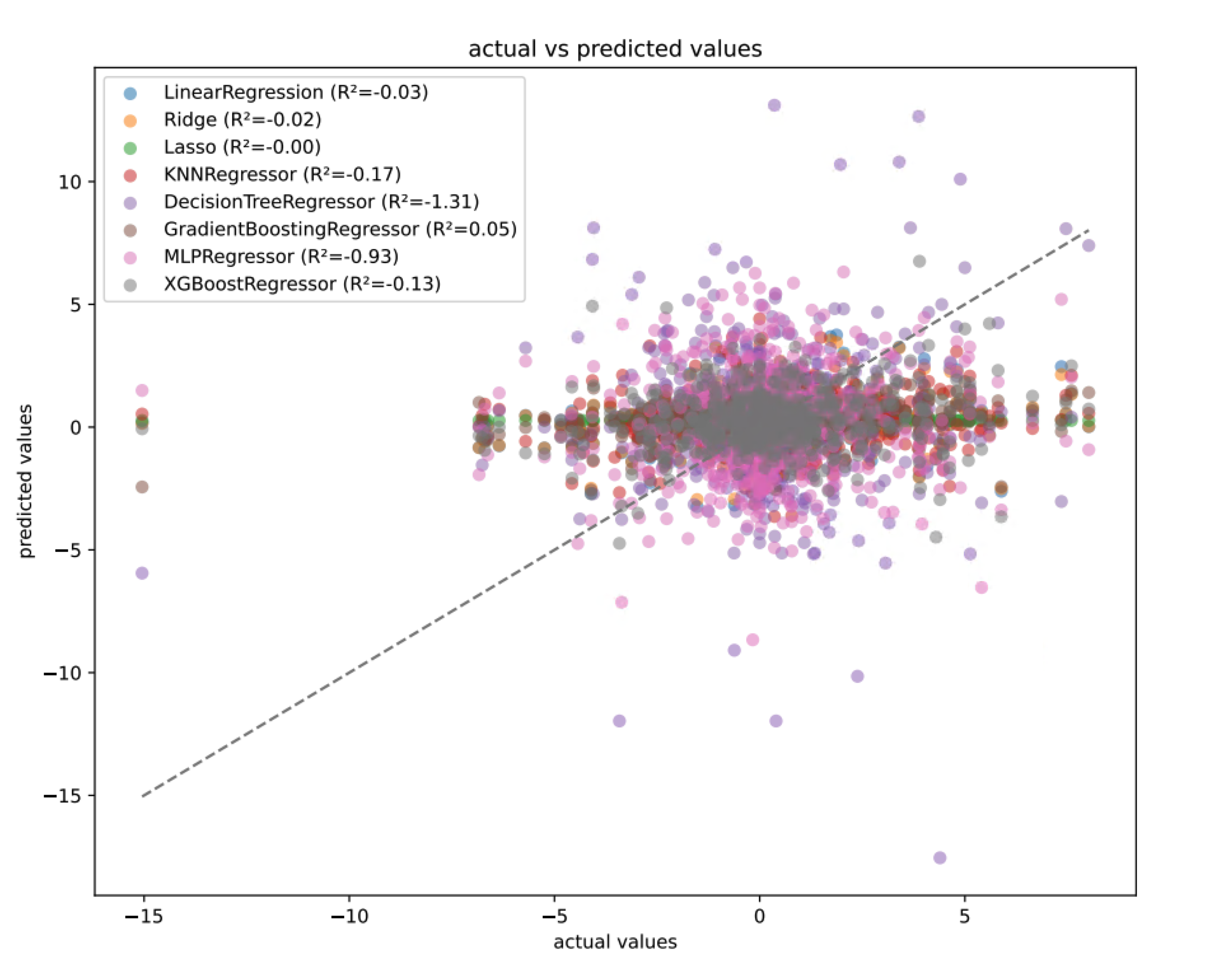

**Supplementary Figure 3: Machine Learning model outcome for the prediction of increase or decrease/no change of BMI in the follow-up visit for each patient.** Comparative machine learning model performance for predicting BMI changes in patients taking olanzapine, presented in two parts with distinct data subsets. The top part examines earliest BMI on olanzapine to follow-up, with (A) ROC curves comparing five models (Logistic Regression, Random Forest, Decision Tree, Gradient Boosting, XGBoost) showing AUC values for the best performing model to ~0.58, and (B) displaying the corresponding Precision-Recall curves. The bottom part analyzes latest BMI on olanzapine to follow-up, (C) with ROC curves for the same five models with similar performance ~0.57, and (D) presenting the associated Precision-Recall curves. The comparison demonstrates that models generally perform a bit better when using the earliest BMI measurements after olanzapine treatment start rather than latest, while GradientBoosting performs similarly across both datasets.

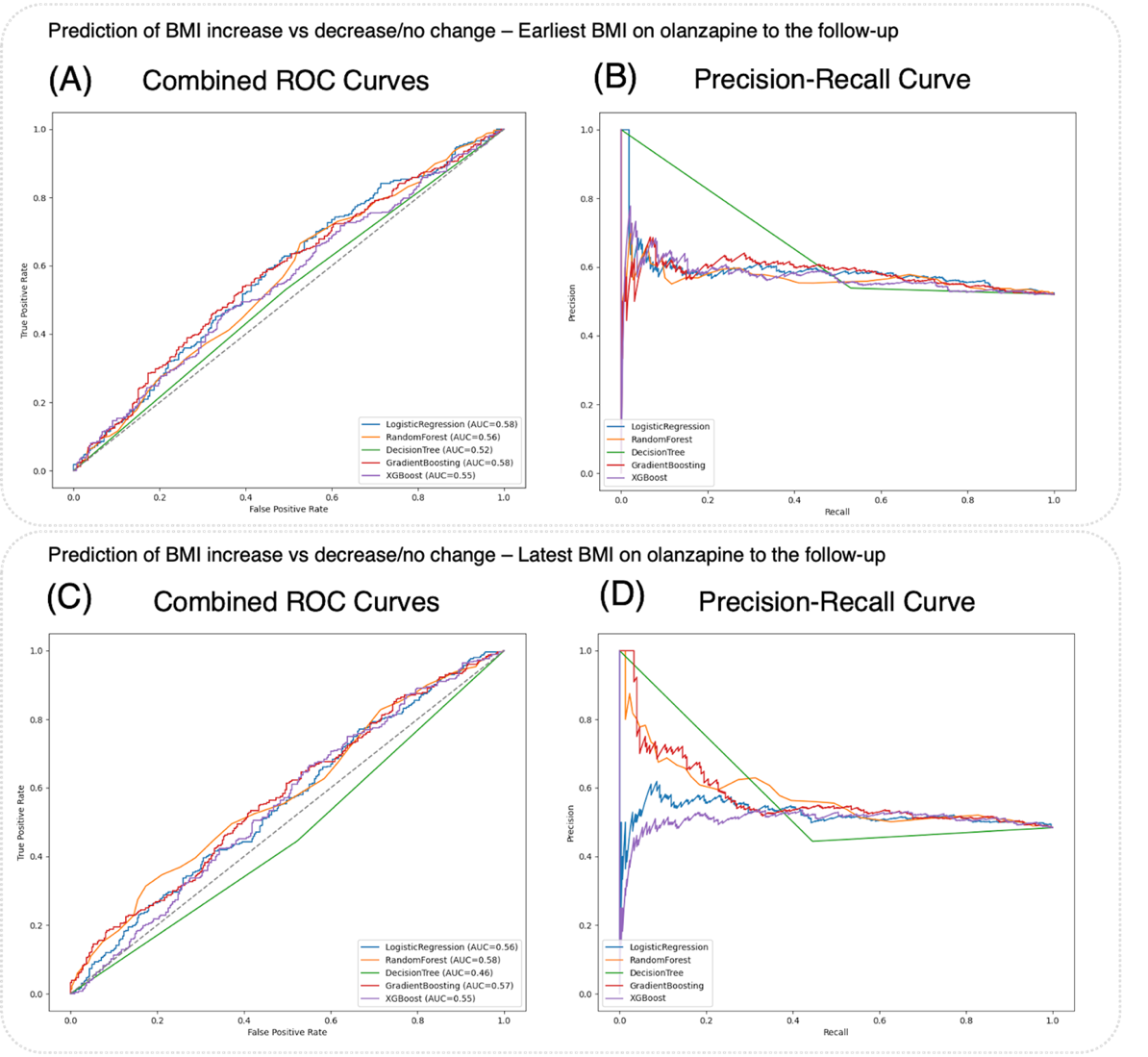

**Supplementary Figure 4. Distributions of BMI-olanzapine related temporal features for individuals treated with olanzapine.** (A, B) Distributions of observations for the earliest BMI records per individual, while (C) and (D) show the latest BMI records per individual on olanzapine. (A, C) Time distance (in days) from start of olanzapine treatment to the first or last BMI measurement while the patients are still taking the medication. (B, D) Time interval (in days) until the next BMI assessment follow-up. (E) Distribution of total number of BMI measurements per individual. Red dashed lines indicate pseudo medians (of 5 values centered around the median) to ensure data anonymity and robustness to outliers. Early BMI measurements after olanzapine initiation typically occur within ~112 days (A), while the latest measurements are around 284 days (C), indicating that in early observations most patients are assessed less than 4 months after medication start while the latest BMI assessment for the majority it is more than 8 months after initiation. The time between measurements also increases from the early (median 142 days in B) to the later follow-up period (median 214 days in D), suggesting reduced monitoring intensity over time. Most patients had a median of 5 BMI measurements during their clinical history (E), though some were monitored much more frequently.

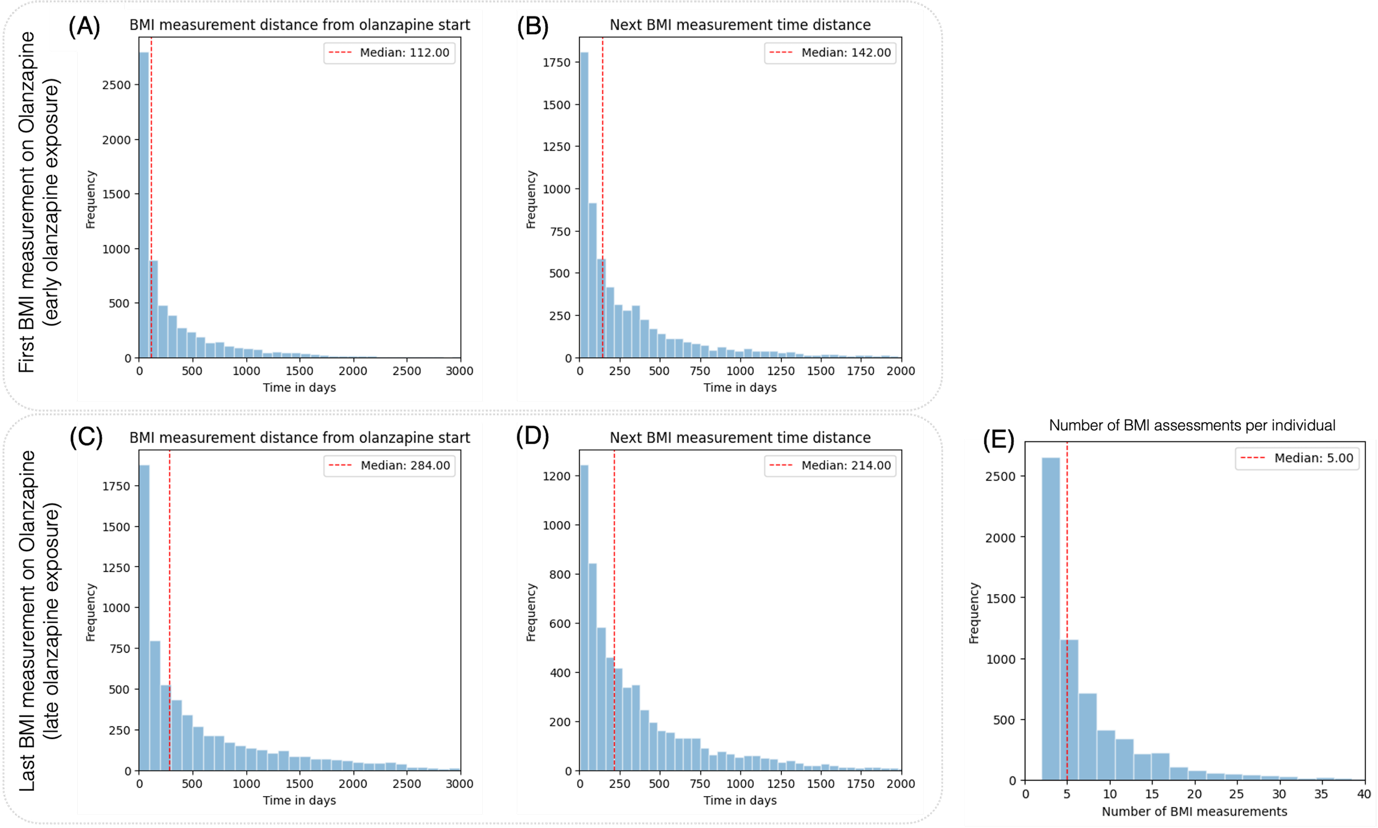

**Supplementary Methods**

The data tables from the registry EHR data included in the study were: BMI, patient demographics, medications, diagnoses, lab tests, hospitalizations and family history. As a first step for the data preprocessing, we ensured that each column in each of the tables in our dataset had the correct type—converting numeric fields to numerical values, treating date fields as proper dates, designating text fields as categorical data, and converting binary indicators to true/false values. We also standardized each table for the handling of missing data by replacing various placeholder strings with a uniform marker for missing entries and removed duplicated records. Finally, we saved the cleaned and type-consistent data tables back into a column-oriented parquet format.

To avoid invalid medication periods and errors besides checking the medication ATC codes to be valid we first filtered the medication data to keep only records with expected dosage ranges for olanzapine (2.5 – 40 mg). Next, we standardized the medication frequency descriptions by mapping irregular text into structured categories, which allowed us to compute a corrected frequency and the corresponding corrected daily dosage. After that, we adjusted the medication intervals by sorting the data, grouping by patient id and medication, and iteratively merging intervals that were less than 48 hours apart—this involved tagging overlapping intervals, aggregating duration and dosage values, and computing a weighted mean for the daily dosage for the aggregation of intervals. As a last step, we enriched the medications dataset by computing additional features: we tracked the cumulative number of unique medications prescribed over time per patient, counted how many times patients had taken specific medications before each record, and determined overlapping medication intervals by processing start and end events, making sure to filter out cases where the end date is before start for a medication.

To prepare the diagnosis data, we iterated over the diagnosis records, and for those missing a valid SKS code, we cleaned the description and tried to extract a valid code using a pattern match. If a direct pattern match failed, we employed a fuzzy matching process with a preset confidence threshold against the official code texts to find the best alternative. Once a code was identified, we updated the record accordingly, cases where no code could be identified were excluded. Next, we shortened the confirmed codes (to reduce sparsity – low frequency of some diagnoses) into a standardized four-digit format (based on the official SKS code hierarchy) and categorized them for streamlined analysis. We then computed the frequency of each diagnosis category per patient by grouping and counting occurrences and then merged these counts back into our dataset. Afterward, we addressed the diagnosis time intervals: for each patient and code, we iteratively merged overlapping intervals by checking if a new diagnosis started within 48 hours of the previous interval's end. This process involved sorting records, comparing current and previous interval dates, tagging overlapping intervals, grouping them, and summing their durations until no further merging was needed.

To calculate BMI changes from one measurement to the next for each patient we first filtered the dataset to keep only adult patients based on their computed age at the time of each BMI assesment. Next, we recalculated BMI values using each patient’s median height, ensuring that only patients with valid height ranges were retained, and removed any extreme BMI values by filtering out the lowest and highest 0.001 percentiles. We also filtered out patients who did not have a sufficient number of BMI measurements to compute any BMI changes. After cleaning the data, we merged the health assessment data with patient demographics records, sorted and de-duplicated the merged data, and aggregated it by patient and BMI measurement timestamp. We then computed BMI trajectories by calculating the differences between consecutive measurements (both prior and subsequent differences), as well as additional metrics like variance in BMI over time and cumulative changes from the first measurement at each BMI timestamp. In addition, we encoded categorical variables such as sex and smoking status into standardized groups, derived a classification for BMI change (indicating weight gain or loss), and computed the rate of BMI change over time by dividing the BMI difference from each measurement to the next with the time elapsing between the measurements.

To determine which types of medications each patient was receiving at the time of each BMI measurement, we merged the medications with the BMI data by mapping medication intervals described by start/end timestamps to patient BMI timestamps—this was achieved through building an interval tree for each patient and applying this mapping in parallel to efficiently attach the appropriate medication codes to each health assessment record. We then we combined the diagnosis data with the BMI timestamps in a similar manner: after filtering and ensuring valid time intervals, we mapped diagnosis codes to patients’ BMI timestamps using the same interval-based approach with parallel processing. The lab results data was then preprocessed by filtering for the relevant patients, cleaning and pivoting the lab test flags so that each test became a separate feature column and aggregating these results by patient and test collection timestamp. We merged these lab results with the BMI data, retaining only those tests performed before the BMI recording time. Following that, we enriched the dataset by merging hospital admission records. This involved filtering admissions for relevant patients, determining whether each patient was currently hospitalized at the time of each BMI timestamp, and calculating total hospitalization counts and durations until that timestamp. We then incorporated family history by transforming the family diagnosis categories in a one-hot encoded format, merging them to the patient-level dataset in the end.

In the next processing stages, we refined the merged dataset by removing sparse columns, standardizing missing values and truncating detailed codes to a four-character format before summarizing them to mitigate sparsity. We filtered for the top 200 most frequent medication and diagnosis codes to be used for downstream modeling. After cleaning up unused columns and ensuring consistency of variable types, we saved the final dataset for further analysis.

We then filtered the data and only kept in the following analysis individuals that received olanzapine and for whom we have BMI reports while they are treated. We also engineered additional features to estimate the number of days a patient was on olanzapine by combining the time differences from the medication timestamps and the duration of medication use. Cases with ambiguous measurement timing relative to the start of medication were excluded. We selected either the first or last BMI measurement for each patient on olanzapine before saving two final dataset tables—one based on the first available record and the other on the last for each individual . Finally, we standardized the target output variable for our 3 different approaches one which was the continuous numerical value of BMI change to follow-up, one to predict a binary outcome for patient BMI gain or loss/0 and finally a binary for >5% BMI increase in the follow-up and —thus completing the data preparation process for further analysis.

To statistically compare BMI between groups of individuals treated with olanzapine and without we checked whether each group follows a normal distribution using a normality test; and since the BMI distributions were skewed and not normal failing the normality test, we applied a nonparametric Mann-Whitney U test, along with computing an approximate z-score. We visualize the data with distribution plots and boxplots are created to visualize BMI differences between treated and untreated groups while annotating statistical significance. We also generate hierarchical clustering heatmaps of Spearman correlations and summarized all the variables reporting pseudo-statistics (averaging groups of 5 extreme and median values) for each numeric column to reduce the risk of re-identification. Building on this, we summarize entire data tables by stratifying them by demographic groups (e.g., sex and age) and calculate appropriate descriptive statistics depending on the column type.

For the regression model data were stratified by unique patient identifiers and binned target values (using quantiles or equal-width cuts) to ensure that splits into training, validation, and test sets maintained a representative distribution of the regression outcome. Once the data was split, we preprocessed it by imputing missing numerical values (using medians), scaling them, and one-hot encoding categorical features whenever it was necessary. After generating the transformed datasets and we trained a suite of regression models (LinearRegression, Ridge, Lasso, KNNRegressor, DecisionTreeRegressor, GradientBoostingRegressor, MLPRegressor, XGBoostRegressor) by fitting each model on the training data and evaluating performance on the test set using metrics like R², mean absolute error, and mean squared error.

For the binary classification modelling we used scikit-learn and complementary numpy, pandas and xgboost library. Data were partitioned into training, and test sets in a stratified fashion by selecting one record per patient and ensuring a 20% allocation for the test set. Preprocessing was accomplished by imputing missing numeric values with the median, scaling them (StandardScaler), and one-hot encoding categorical variables (imputing missing entries with the most frequent value). The classification models evaluated, included Logistic Regression (with max_iter=5000), Random Forest, Decision Tree, Gradient Boosting, and XGBoost (configured with objective 'binary:logistic'). To address class imbalance, the decision threshold in each model was optimised based on the optimal F1 score for the positive class. Visualization of performance included confusion matrices, AUROC curves as specified by the configuration parameters. For hyperparameter tuning, 10 fold cross validation was applied and parameters were optimised based on validation AUROC using predefined parameter grids (e.g., Logistic Regression with C values of 0.1, 1, and 10 using lbfgs solver; Random Forest with n_estimators of 100 and 200 and max_depth of None, 10, and 20) on a combined training/validation set via a PredefinedSplit. Model interpretability was enhanced using SHAP: tree-based models were explained with TreeExplainer (with interventional feature perturbation), Logistic Regression with LinearExplainer (using a background sample of 100 observations), and other models with KernelExplainer; SHAP summary plots (layered violin plots of the top 15 features. In addition, subgroup analysis was performed by stratifying patients by age—using bins of age groups 18–29, 30–49, 50–69, and 70+ and by sex and subgroup-specific ROC curves were computed to assess model performance across different demographic groups. The entire pipeline, was performed in Python (Python version 3.13).
